# Clinical Characteristics of Children with COVID-19: A Rapid Review and Meta-Analysis

**DOI:** 10.1101/2020.04.13.20064352

**Authors:** Zijun Wang, Qi Zhou, Chenglin Wang, Qianling Shi, Shuya Lu, Yanfang Ma, Xufei Luo, Yangqin Xun, Weiguo Li, Muna Baskota, Yinmei Yang, Hui Zhai, Toshio Fukuoka, Hyeong Sik Ahn, Myeong Soo Lee, Zhengxiu Luo, Enmei Liu, Yaolong Chen, on behalf of COVID-19 evidence and recommendations working group

**Affiliations:** Evidence-based Medicine Center, School of Basic Medical Sciences, Lanzhou University, Lanzhou 730000, China; The First School of Clinical Medicine, Lanzhou University, Lanzhou 730000, China; Children’s Hospital of Chongqing Medical University, Chongqing 400014, China; Department of Pediatric, Sichuan Provincial People’s Hospital, University of Electronic Science and Technology of China, Chengdu 611731, China; Chinese Academy of Sciences Sichuan Translational Medicine Research Hospital, Chengdu 610072, China; School of Public Health, Lanzhou University, Lanzhou 730000, China; Department of Respiratory Medicine, Children’s Hospital of Chongqing Medical University, Chongqing 400014, China; National Clinical Research Center for Child Health and Diseases, Ministry of Education Key Laboratory of Child Development and Disorders, China International Science and Technology Cooperation Base of Child Development and Critical Disorders, Children’s Hospital of Chongqing Medical University, Chongqing 400014, China; Chongqing Key Laboratory of Pediatrics, Chongqing 400014, China; Emergency and Critical Care Center, the Department of General Medicine, Department of Research and Medical Education at Kurashiki Central Hospital, Japan; Advisory Committee in Cochrane Japan, Japan; Department of Preventive Medicine, Korea University College of Medicine, Seoul, Korea; Korea Cochrane Centre, Korea; Korea Institute of Oriental Medicine, Daejeon, Korea; University of Science and Technology, Daejeon, Korea; Lanzhou University, an Affiliate of the Cochrane China Network, Lanzhou 730000, China; Chinese GRADE Center, Lanzhou 730000, China; Key Laboratory of Evidence Based Medicine and Knowledge Translation of Gansu Province, Lanzhou University, Lanzhou 730000, China

**Author notes:** These authors contributed equally to this work (Zijun Wang, Qi Zhou, Chenglin Wang). **Correspondence to:** Enmei Liu. Department of Respiratory Medicine, Children’s Hospital of Chongqing Medical University, Chongqing 400014, China.; Yaolong Chen. Evidence-based Medicine Center, School of Basic Medical Sciences, Lanzhou University, Lanzhou 730000, China.

**Keywords:** Children, clinical characteristics, COVID-19, meta-analysis, rapid review

## Abstract

**Background:** Most guidelines on COVID-19 published so far include recommendations for patients regardless of age. Clinicians need a more accurate understanding of the clinical characteristics of children with COVID-19.

**Methods:** We searched studies reporting clinical characteristics in children with COVID-19 published until March 31, 2020. We screened the literature, extracted the data and evaluated the risk of bias and quality of evidence of the included studies. We combined some of the outcomes (symptoms) in a single-arm meta-analysis using a random-effects model.

**Results:** Our search retrieved 49 studies, including 25 case reports, 23 case series and one cohort study, with a total of 1667 patients. Our meta-analysis showed that most children with COVID-19 have mild symptoms. Eighty-three percent of the children were within family clusters of cases, and 19% had no symptoms. At least 7% with digestive symptoms. The main symptoms of children were fever (48%, 95% confidence interval [CI]: 39%, 56%) and cough (39%, 95% CI: 30%, 48%). The lymphocyte count was below normal level in only 15% [95% CI: 8%, 22%] of children which is different from adult patients. 66% [95% CI: 55%, 77%] of children had abnormal findings in CT imaging.

**Conclusions:** Most children with COVID-19 have only mild symptoms, and many children are asymptomatic. Fever and cough are the most common symptoms in children. Vomiting and diarrhea were not common in children. The lymphocyte count is usually within the normal range in children.

## Background

In December 2019, a previously unknown type of pneumonia broke out in Wuhan, China, which was later confirmed to be caused by a novel type of beta coronavirus, Severe Acute Respiratory Syndrome Coronavirus 2 (SARS-CoV-2). In February 2020, the World Health Organization (WHO) officially named the disease as “Coronavirus Disease 2019 (COVID-19)” (1). Like MERS-CoV and SARS-CoV, SARS-CoV-2 can also be transmitted between humans (2-5). Since the first occurrence of COVID-19 case in Wuhan, Hubei Province, China on December 8, 2019 (6, 7), the disease is spreading rapidly. WHO reassessed the potential impact of COVID-19 on global public health and subsequently declared COVID-19 as Public Health Emergency of International Concern (PHEIC) on January 30, 2020.

Research has proven that people of all ages are susceptible to SARS-CoV-2. The mean age of COVID-19 patients was 47 years, with 55% of the patients being between 15 and 49 years old. Only 9% of the patients were under 15 years old.(8) For this reason, most of the guidelines published so far include recommendations for patients regardless of age, only a few recommendations are for children. Although the great majority of patients are adults, children’s respiratory structural characteristics and immune response system differ essentially from those in adult (9-11), and the diagnostic criteria and management according to recommendations targeting adults may not be appropriate for children. Our study aims therefore to identify the clinical features of children with COVID-19, help clinicians to confirm and treat the suspected children as soon as possible, and provide support for the development of guidelines for COVID-19 in children.

## Methods

### Search strategy

We comprehensively searched the following electronic databases: Cochrane library, MEDLINE (via PubMed), Embase, Web of Science, China Biology Medicine disc (CBM), China National Knowledge Infrastructure (CNKI), and Wanfang Data from their inception until March 31, 2020 with the terms “2019-novel coronavirus”, “SARS-CoV-2”, “COVID-19”, “2019-nCoV”, “clinical features” and their derivatives. We also searched World Health Organization (WHO), Chinese Center for Disease Control and Prevention (CCDC), National Health Commission of the People’s Republic of China, USA National Institutes of Health Ongoing Trials Register (ClinicalTrials.gov), the ISRCTN registry, Google Scholar and the preprint servers medRxiv (https://www.medrxiv.org/), bioRxiv (https://www.biorxiv.org/) and SSRN (https://www.ssrn.com/index.cfm/en/). In addition, we searched the reference lists of the identified studies for further potential studies. The full search strategy can be found in *Supplementary Material 1*.

### Inclusion and exclusion criteria

We included studies on children (aged <18 years) with COVID-19 that report clinical features of patients, such as symptoms, signs, laboratory examinations and imaging manifestations. Diagnosis of COVID-19 was based on the Novel Coronavirus Pneumonia Prevention and Control Program (7th edition) issued by the National Health and Health Committee of China (12) and surveillance case definitions for human infection with novel coronavirus (nCoV) Interim guidance v2 issued by World Health Organization (13).We excluded in vitro studies, Traditional Chinese Medicine studies, conference abstracts, comments, letters, and duplicates, and studies where we could not extract the data. We made no restrictions on language and publication status.

### Study selection

Two reviewers (Z Wang and C Wang) selected the studies independently after first eliminating duplicates. The bibliographic software EndNote was used and any discrepancies were settled by discussion, consulting a third reviewer (Q Zhou) if necessary. Before the formal selection, the reviewers searched a random sample of 50 citations. The reviewers screened first all titles and abstracts with the pre-defined criteria, and categorized the articles into three (eligible, not eligible, and unclear) groups. In the second Step, full-texts of those potentially eligible or unclear studies were reviewed to identify the final inclusion. All the reasons for exclusion of ineligible studies were recorded, and the process of study selection was documented using a PRISMA flow diagram (14, 15).

### Data extraction

Two reviewers (Q Shi and S Lu) extracted the data independently with a standardized data collection form, including:1) basic information (e.g. first author); 2) symptoms; 3) routine blood tests (e.g. leucocyte count); 4) blood biochemistry (e.g. alanine aminotransferase); 5) coagulation function (e.g. activated partial thromboplastin time); 6) imaging findings (e.g. abnormal imaging). For dichotomous outcomes, we abstracted the number of events and total participants per group. For continuous outcomes, we abstracted means, standard deviations (SD), and the number of total participants in per group. Outcomes with no events were reported, but these were excluded from the meta-analysis. If means and standard deviations (SD) were not reported, we calculated them from the reported indicators. (16) If data were missing or reported in an unusable way, we excluded the study from the meta-analysis and report the findings descriptively.

### Risk of bias assessment

Two reviewers (Z Wang and C Wang) assess the risk of bias in each study independently. Discrepancies were settled by discussion, consulting a third reviewer (Q Zhou) if necessary. For randomized controlled trials (RCTs), we will assess the risk of bias independently using Cochrane risk-of-bias tool (17). It consists of seven domains, for each, we will grade as “Low”, “Unclear”, and “High”. For nonrandomized controlled trials (nRCTs), ROBINS-I tool will be used(18). It consists of seven domains, for each, we will grade as “Low risk”, “Moderate risk”, “Serious risk”, “Critical risk”, and “No information”. For case-control and cohort studies, the Newcastle-Ottawa Scale will be used (19, 20). It consists of eight domains, for each, we will grade with stars. The more stars, the lower the risk of bias. For cross-sectional studies, we use a methodology evaluation tool recommended by Agency for Healthcare Research and Quality (AHRQ) (21). This tool assesses the quality of bias according to 11 criteria. And each criterion is answered by “Yes”, “No” or “unsure”. For case reports and case series, we used a methodology evaluation tool recommended by National Institute for Health and Care Excellence (NICE) (22). The risk of bias is evaluated according to eight criteria. The results were summarized by scoring method, for the “Yes” items, the score was 1, and for the “no” items, the score was 0. The higher the total score, the lower the risk of bias.

### Data synthesis

We summarized the results of the studies including less than 9 patients and did meta-analysis of included studies that have at least 9 patients. For dichotomous outcomes, we did a meta-analysis of proportions, reporting the effect size (ES) with 95% confidence intervals (CI). For continuous outcomes, we did a meta-analysis of continuous variable, calculating the effect size (ES) with 95% CI. We described the results of the studies with patients that below nine. As clinical and methodological heterogeneity in the study design, characteristics of participants, interventions and outcome measures was expected, we used random-effects models (23). Two-sided *P* values < 0.05 were considered statistically significant. Heterogeneity was defined as *P* values <0.10 and *I*^*2*^>50%. All analyses were performed in STATA version 14.

### Quality of the evidence assessment

Two reviewers (Q Zhou and Y Xun) assessed the quality of main evidence independently using the Grading of Recommendations Assessment, Development and Evaluation (GRADE) tool. We produced a “Summary of Findings” table using the GRADE pro software (24, 25). Direct evidence from RCTs is first set as high quality, and evidence from observational studies as low quality. Then initial quality can then be downgraded for five reasons (study limitations, consistency of effect, imprecision, indirectness, and publication bias) and upgraded for three reasons (large magnitude of effect, dose-response relation and plausible confounders or biases) (26-31). Finally, the quality of main evidence can be classified as high, moderate, low, or very low, which reflects the extent to what we can be confident that the effect estimates are correct.

As COVID-19 is a public health emergency of international concern and the situation is evolving rapidly, our study was not registered in order to speed up the process. (32)

## Results

### Study selection and characteristics

Our initial search retrieved 774 records (*Figure 1*). After removing duplicates, we screened the titles, abstracts and full texts, and 49 studies were finally included. The articles included 25 case reports, 23 case series and one cohort study. The studies included a total of 1667 patients: 955 males and 712 females. Eighteen percent of children were aged less than one year. Most of the studies were carried out in China, including 17 studies from Hubei (*Table 1*). One study was from Singapore, one from Korea, one from Vietnam and one from Iran. The results on risk of bias are reported in *Supplementary Material 2*, and quality of the evidence in *Supplementary Material 3*.

**Table 1.** Characteristics of included studies.

| First author | Study Location | Number |  |  | Age (year) | Study design |
| --- | --- | --- | --- | --- | --- | --- |
|  |  | All | M | F |  |  |
| Deng 2020 (33) | Xian | 2 | 1 | 1 | 8.0±7.1 | case report |
| Cai 2020 (34) | Shanghai | 1 | 1 | 0 | 7 | case report |
| Zhang 2020 (35) | Hunan | 1 | 0 | 1 | 0.2 (0.1, 0.3) | case report |
| Wei 2020 (36) | Hubei | 9 | 2 | 7 | 0.58 (0.28, 0.79) | case series |
| Chen 2020 (37) | Hubei | 1 | 1 | 0 | 1.1 | case report |
| Zeng 2020 (38) | Hubei | 1 | 1 | 0 | 0 | case report |
| Feng 2020 (39) | Shenzhen | 15 | 5 | 10 | 8.0±2.9 | case series |
| Wang 2020 (40) | Hubei | 1 | 1 | 0 | 5.6 | case report |
| Quan 2020 (41) | Liaoning | 1 | 0 | 1 | 4 | case report |
| Xiong 2020 (42) | Chongqing | 2 | 1 | 1 | 5.2/5.2 | case report |
| Cai 2020 (43) | Shanghai | 10 | 4 | 6 | 6.5 (4.0, 9.0) | case series |
| Wang 2020 (44) | Xian | 31 | 15 | 16 | 7.9±4.0 | case series |
| Tang 2020 (45) | Shenzhen | 26 | 9 | 17 | 6.90±0.70 | case series |
| Liu 2020 (46) | Hubei | 6 | 2 | 4 | 3.0 (3.0, 4.0) | case report |
| Lu 2020 (47) | Hubei | 171 | 104 | 67 | 7.1±2.8 | case series |
| Ma 2020 (48) | Hubei | 50 | 28 | 22 | 2.5 (0.9, 7.0) | case series |
| Zhao 2020 (49) | Zhejiang | 1 | 1 | 0 | 13 | case report |
| Li 2020 (50) | Hubei | 30 | 18 | 12 | 6.5±3.4 | case series |
| Zhang 2020 (51) | Shandong | 10 | 3 | 7 | 5.7±4.2 | case series |
| Zhang 2020 (52) | Hunan | 2 | 0 | 2 | 1.2/1.2 | case report |
| Liu 2020 (53) | Hubei | 1 | 1 | 0 | 9 | case report |
| Xiao 2020 (54) | Chongqing | 1 | 0 | 1 | 7.83 | case report |
| Xu 2020 (55) | Chongqing | 32 | 17 | 15 | 9.0±4.7 | case series |
| Chan 2020 (56) | Hong Kong | 1 | 1 | 0 | 10 | case report |
| Kam 2020 (57) | Singapore | 1 | 1 | 0 | 0.5 | case report |
| Park 2020 (58) | Korea | 1 | 0 | 1 | 10 | case report |
| Du 2020 (59) | Shandong | 14 | 6 | 8 | 7.1±4.7 | case series |
| Liu 2020 (60) | Shanghai | 4 | 2 | 2 | 3.8±4.1 | case report |
| Wang 2020 (61) | Hubei | 1 | 1 | 0 | 0 | case report |
| Rahimzadeh 2020 (62) | Iranian | 9 | 6 | 3 | 5.0 (3.5, 5.5) | case series |
| Ji 2020 (63) | Beijing | 2 | 2 | 0 | 9/15 | case report |
| Xu 2020 (64) | Guangzhou | 10 | 6 | 4 | 6.6 (1.8, 13.7) | case series |
| Xia 2020 (65) | Hubei | 20 | 13 | 7 | 4.7±3.9 | case series |
| Dong 2020 (66) | Shanghai | 731 | 420 | 311 | 10 (NR) | case series |
| Xing 2020 (67) | Hong Kong | 3 | 2 | 1 | 1.5/2.5/3.6 | case report |
| Yu 2020 (68) | Hubei | 82 | 51 | 31 | NR (0-16) | case series |
| Liu 2020 (69) | Hubei | 91 | 56 | 35 | NA | case series |
| Ma 2020 (70) | Hubei | 115 | 73 | 42 | NA | case series |
| Qian 2020 (71) | Zhejiang | 2 | 0 | 2 | 1.1 | case report |
| Qiu 2020 (72) | Zhejiang | 36 | 23 | 13 | 8.3±3.5 | cohort study |
| Zhang 2020 (73) | Hubei | 25 | 14 | 11 | 3.0 (2.0, 9.0). | case series |
| Sun 2020 (74) | Hubei | 8 | 6 | 2 | 5.0 (1.0, 13.7) | case report |
| Dong 2020 (75) | Hubei | 2 | 2 | 0 | 3/2.3 | case report |
| Wu 2020 (76) | Shandong | 74 | 44 | 30 | 6.0 (0.1, 15.1) | case series |
| Su 2020 (77) | Shandong | 9 | 3 | 6 | 3.6 (1.8, 6.8) | case series |
| Le 2020 (78) | Vietnam | 1 | 0 | 1 | 0.3 | case report |
| Zhong 2020 (79) | Hunan | 9 | 4 | 5 | 8 (1.5, 10.0) | case series |
| Yang 2020 (80) | Shandong | 10 | 3 | 7 | 4 (2, 8) | case series |
| Tang 2020 (81) | Zhejiang | 1 | 1 | 0 | 10 | case report |

Ages were reported either as mean ± standard deviation, or median (interquartile range), or single year. NR: not report; NA: not applicable.

**Figure 1.**
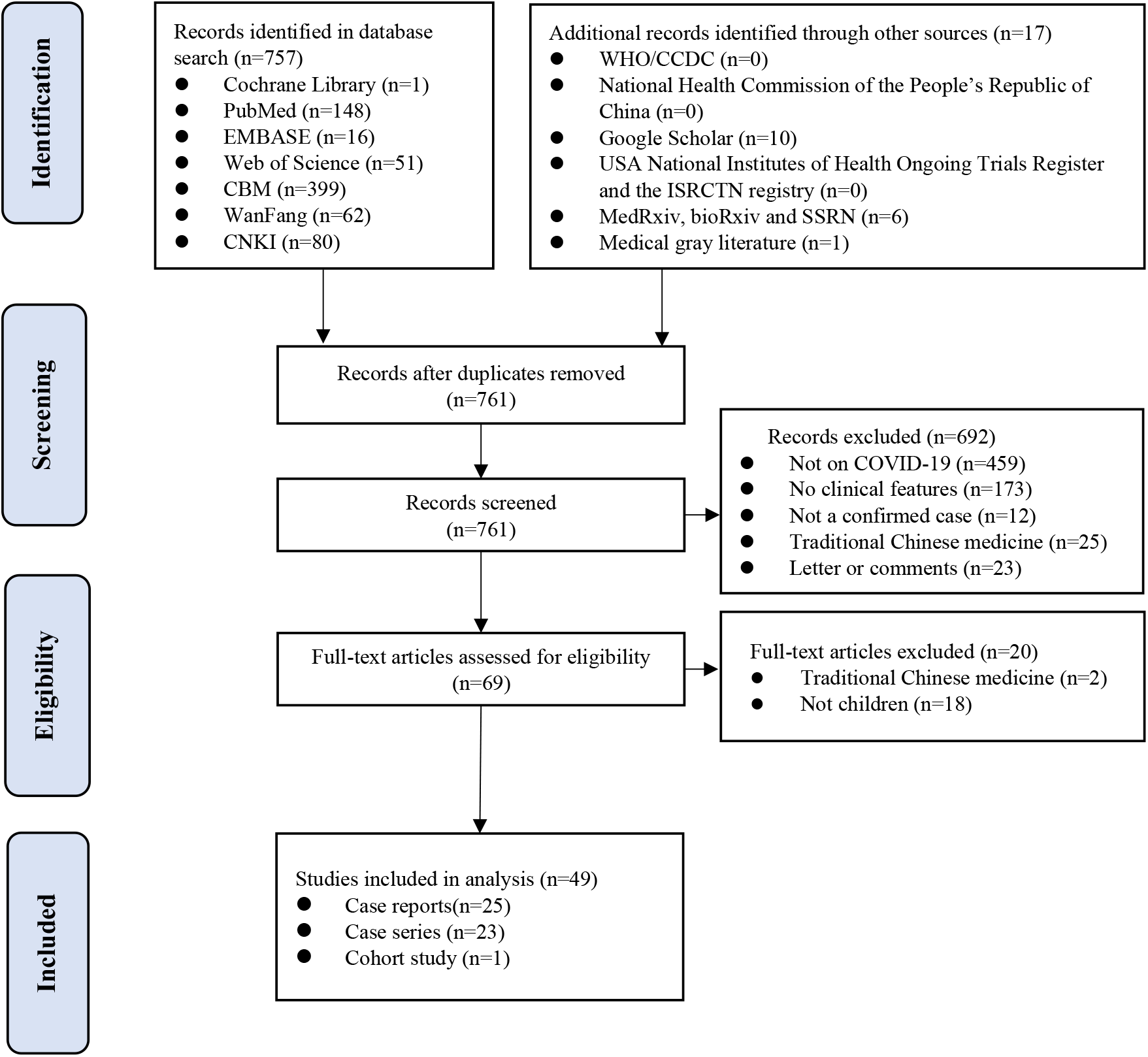
Flow diagram.

### Symptoms and imaging results

All of the included studies reported the symptoms of children with COVID-19. The results showed that 83% [95% CI: 78%, 88%] of the cases had likely acquired the infection from their family members with COVID-19. 94% [95% CI: 90%, 98%] of children were mild cases and 3% [95% CI: 2%, 4%] were severe case. Among the children with severe symptoms that reported symptom clearly, 9 children have comorbidity, 10 children have gastrointestinal symptoms and 4 children have concurrent infection. Only two children were dead that has been reported in our included studies.(47, 66) The main symptoms were fever (48% [95% CI: 39%, 56%]), cough (39% [95% CI: 30%, 48%]). Thirty percent [95% CI: 18%, 42%] of children had both cough and fever. 7% [95% CI: 5%, 9%] and 6% [95% CI: 4%, 9%] of cases had diarrhea and nausea/vomiting. The proportion of children with more than one symptom was 35% [95% CI: 21%, 48%], and 19% [95% CI: 14%, 23%] of all children were asymptomatic.

Forty-two studies reported the imaging features of children with COVID-19, including 19 case series and 23 case reports. 66% [95% CI: 55%, 77%] had abnormal imaging. 35% [95% CI: 26%, 44%] of children had ground-glass opacity.

More information can be found in *Table 2* and *Supplementary Material 4*.

**Table 2.** Symptoms and imaging results of patients with COVID-19.

| The overall symptoms | ES | $I^2$ | P-value |
| --- | --- | --- | --- |
| No Symptom | 19% (14%, 23%) | 70.7% | <0.10 |
| Mild symptom | 94% (90%, 98%) | 89.9% | <0.10 |
| Severe symptom | 3% (2%, 4%) | 22.5% | 0.24 |
| More than one sign or symptom | 35% (21%, 48%) | 80.8% | <0.10 |
| Specific symptoms |  |  |  |
| Fever | 48% (39%, 56%) | 85.7% | <0.10 |
| Cough | 39% (30%, 48%) | 84.4% | <0.10 |
| Fever and Cough | 30% (18%, 42%) | 67.4% | <0.10 |
| Sputum production | 19% (0%, 44%) | 97.5% | <0.10 |
| Rhinorrhoea | 9% (6%, 12%) | 0.0% | 0.79 |
| Shortness of breath / Dyspnoea | 9% (0%, 19%) | 91.8% | <0.10 |
| Myalgia or fatigue | 8% (5%, 12%) | 56.7% | <0.10 |
| Diarrhoea | 7% (5%, 9%) | 0.0% | 0.62 |
| Nausea or vomiting | 6% (4%, 9%) | 0.0% | 1.00 |
| Nasal obstruction | 6% (3%, 9%) | 0.0% | 0.59 |
| Sore throat | 6% (2%, 10%) | 35.2% | 0.13 |
| Headache | 4% (1%, 6%) | 0.0% | 0.61 |
| Imaging findings |  |  |  |
| Abnormal | 66% (55%, 77%) | 89.6% | <0.10 |
| Unilateral pneumonia | 31% (20%, 43%) | 81.5% | <0.10 |
| Bilateral pneumonia | 28% (20%, 36%) | 81.3% | <0.10 |
| Ground-glass opacity | 35% (26%, 44%) | 84.7% | <0.10 |

### Laboratory results

Seventeen case series reported the results of routine blood tests. The mean leucocyte count in children was 6.25 ×10 ^9^/L [95% CI: 5.97, 6.54]. 15% [95%CI: 4%, 26%] of cases had leucocyte count above the normal range and 28% [95% CI: 17%, 38%] of cases below the normal range. The mean lymphocyte count in children was 2.84×10 ^9^/L [95% CI: 2.55, 3.13]. Lymphocyte count was elevated in 41% [95%CI: 2%, 80%] and below normal in 15% [95% CI: 8%, 22%] of children.

Eighteen case series reported the results of blood biochemistry tests. The mean value of alanine aminotransferase (ALT) was 20.46 U/L [95% CI: 14.51, 26.41], and 11% [95% CI: 8%, 14%] of cases had elevated ALT values. The mean value of aspartate aminotransferase (AST) was 32.04 U/L [95% CI: 30.25, 33.83], and 15% [95% CI: 9%, 21%] of cases had elevated AST values. The mean value of C-Reactive Protein (CRP) was 5.05 mg/L [95% CI: 1.86, 8.24], and CRP was elevated in 22% [95% CI: 15%, 28%] of the children.

Nine case series reported coagulation function test, the mean value of D-dimer was 0.33mg/L [95% CI: 0.17, 0.49] in studies of children with COVID-19. 15% [95% CI: 7%, 22%] of cases above the normal range of D-dimer value.

More information can be found in *Table 3* and *Supplementary Material 4*.

**Table 3.** Laboratory results of children with COVID-19.

| Routine blood values | ES | $I^2$ | $P$-value |
| --- | --- | --- | --- |
| Leucocytes ($\times 10^9/L$) | 6.25 (5.97, 6.54) | 0.0% | 0.69 |
| Above normal range | 15% (4%, 26%) | 77.3% | <0.10 |
| Below normal range | 28% (17%, 38%) | 85.4% | <0.10 |
| Lymphocytes ($\times 10^9/L$) | 2.84 (2.55, 3.13) | 77.8% | <0.10 |
| Above normal range | 41% (2%, 80%) | 98.5% | <0.10 |
| Below normal range | 15% (8%, 22%) | 79.9% | <0.10 |
| Neutrophils ($\times 10^9/L$) | 2.70 (2.10, 3.31) | 93.8% | <0.10 |
| Above normal range | 23% (0%, 48%) | 85.8% | <0.10 |
| Below normal range | 24% (4%, 44%) | 58.5% | <0.10 |
| Platelets ($\times 10^9/L$) | 264.65 (254.17, 275.12) | 91.2% | <0.10 |
| Above normal range | 10% (3%, 17%) | 45.5% | 0.14 |
| Below normal range | 6% (0%, 16%) | 80.6% | <0.10 |
| Haemoglobin (g/L) | 127.21 (123.38, 131.04) | 97.4% | <0.10 |
| Above normal range | 13% (4%, 22%) | 58.4% | <0.10 |
| Below normal range | 5% (0%, 11%) | 38.5% | 0.20 |
| **Blood biochemistry** |  |  |  |
| Albumin (g/L) | 45.30 (45.13, 45.47) | 0.0% | 0.70 |
| Below normal range | 35% (25%, 45%) | 0.0% | 0.33 |
| Alanine aminotransferase (U/L) | 20.46 (14.51, 26.41) | 96.9% | <0.10 |
| Above normal range | 11% (8%, 14%) | 0.0% | 0.72 |
| Aspartate aminotransferase (U/L) | 32.04 (30.25, 33.83) | 49.2% | <0.10 |
| Above normal range | 15% (9%, 21%) | 45.8% | <0.10 |
| Total bilirubin ($\mu\text{mol/L}$) | 8.14 (1.45, 14.82) | 97.5% | <0.10 |
| Above normal range | 3% (0%, 6%) | NA | NA |
| Blood urea nitrogen (mmol/L) | 3.81 (3.43, 4.18) | 0.0% | 0.56 |
| Above normal range | 12% (0%, 33%) | 94.9% | <0.10 |
| Below normal range | 14% (0%, 35%) | 57.6% | 0.13 |
| Creatinine ($\mu\text{mol/L}$) | 41.60 (32.98, 50.22) | 95.6% | <0.10 |
| Above normal range | 12% (0%, 33%) | 94.9% | <0.10 |
| Below normal range | 8% (3%, 14%) | 0.0% | 0.59 |
| Creatine kinase (U/L) | 104.37 (95.66, 113.08) | 56.6% | 0.10 |
| Above normal range | 13% (0%, 38%) | 77.0% | <0.10 |
| Lactate dehydrogenase (U/L) | 264.43 (241.85, 287.02) | 98.1% | <0.10 |
| Above normal range | 38% (25%, 51%) | 72.9% | <0.10 |
| Below normal range | 11% (0%, 32%) | NA | NA |
| Myoglobin (ug/L) | 15.33 (11.18, 19.48) | NA | NA |
| Procalcitonin (g/L) | 0.06 (0.00, 0.16) | 0.0% | 1.00 |
| Above normal range | 44% (20%, 69%) | 98.0% | <0.10 |
| CRP (mg/L) | 5.05 (1.86, 8.24) | 86.9% | <0.10 |
| Above normal range | 22% (15%, 28%) | 71.3% | <0.10 |
| **Coagulation function** |  |  |  |
| Activated partial thromboplastin time (s) | 37.59 (28.69, 46.48) | 96.4% | <0.10 |
| Increased | 11% (5%, 18%) | 0.0% | 0.72 |
| Decreased | 4% (0%, 8%) | NA | NA |
| Prothrombin time (s) | 12.25 (11.30, 13.20) | 99.0% | <0.10 |
| Increased | 2% (0%, 4%) | 0.0% | 0.42 |
| Decreased | 2% (0%, 6%) | NA | NA |
| D-dimer (mg/L) | 0.33 (0.17, 0.49) | 0.0% | 0.98 |
| Increased | 15% (7%, 22%) | 38.4% | 0.17 |

NA: not applicable

## Discussion

### Principal findings

Children had on average milder symptoms, with many children having even no symptoms. Most children infected with COVID-19 were exposed through family clusters. About half of children present with fever or cough, and about one third of children with both fever and cough. Only a small minority of children had vomiting or diarrhea as initial symptoms. Leucocyte and lymphocyte counts are often in normal or above the normal range in children with COVID-19. Abnormalities in CT imaging were found in more than half of the children, the most common being ground-glass opacity in unilateral lung.

The course of COVID-19 in children can be characterized by mild illness and no symptoms. According to a study by the Chinese CDC, as of February 11, 2020, 81% of all patients with COVID-19 showed only mild symptoms. (82) However, although the disease was less severe in the majority of adults (51%-74%) (83-85), 26%-32% of adults were still committed to ICU, and had often basic diseases such as hypertension or diabetes.(86-88) In contrast, we found only 3% report of children with severe illness. The Chinese CDC also pointed out that 889 (1%) patients with COVID-19 were asymptomatic (82); in our study, about 19% of children were completely asymptomatic, which is higher than the average level of the whole patient significantly. A study of asymptomatic infections with COVID-19 also showed that 29.2% of cases showed normal CT image and had no symptoms during hospitalization. What’s more, these cases were younger (median age: 14.0 years; P=0.012) than the rest.(89) Of children with COVID-19, 83% had other family members infected. The majority of asymptomatic children in family clusters were confirmed after a positive nucleic acid test, which was conducted because of the close contact to infected family members. It seems that family cluster in children were more likely to be tested than adults. So, we suspect whether asymptomatic children are really asymptomatic or are still in the incubation period. Another explanation could be that only symptomatic cases transmit; So, adults without symptoms are seldom diagnosed, while children without symptoms (who had not that many any other contacts during the holiday season than their family members) get diagnosed anyway because of the obvious exposure.

The diagnosis of suspected cases in children needs comprehensive consideration. Fever and cough were the main symptoms in patients with COVID-19, which is reported by the most of current guidelines and recommendations. (12, 90-93) Although fever and cough also the first two symptom of children. When compared with adults, fever and cough occurred only in 48% and 39% of children, respectively. The rates of fever and cough in adults are up to 98%(88) and 87% (84), which indicates that fever and cough in children are not as common as in adults. Compared with children infected with SARS, MERS and other viral diseases (94, 95), there are no specific symptoms in children with COVID-19 that could help to diagnose the disease accurately. Therefore, detection methods are particularly important for the diagnosis of COVID-19. Chinese National Health Commission also pointed out that fever and/or respiratory symptoms, imaging features indicating of pneumonia, leukocyte and lymphocyte counts characteristics in the early stage, and epidemiological history should be comprehensively used to determine suspected cases. After that, RT-PCR, sequencing or specific antibody were used to make a definite diagnosis.(12)

Attention should be paid to the children with COVID-19 who start with gastrointestinal symptoms. Although gastrointestinal symptoms such as nausea, vomiting and diarrhea are less common in children with COVID-19, recent studies have SARS-CoV-2 in the feces of patients (96, 97) and a study showed that some children persistently tested positive on rectal swabs even after nasopharyngeal testing was negative, raising the possibility of fecal–oral transmission.(98) According to one study, diarrhea was the first symptom in three out of 31 children.(88) Moreover, three of the eight children with severe cases of COVID-19 that reported symptom clearly had gastrointestinal symptoms, one of them started with gastrointestinal symptoms, without any obvious respiratory tract infection in the early stage (37). In addition, comorbidity cannot be ignored either. Our results showed that half of these eight children with severe cases had other diseases, including two with intussusception and one of them was dead (47). Similarly, a study of children with MERS also suggest that serious illness can occur in children with underlying disease.(99) Although there is no evidence that gastrointestinal symptoms and comorbidity in children are related to the severity of the disease, clinicians should pay attention to the gastrointestinal symptoms and comorbidity in the process of diagnosing children with COVID-19 and apply real-time monitoring and protection.

Abnormal CT imaging was less common in children with COVID-19 than adults, but the imaging findings were similar in children and adults. Unilateral pneumonia is common in children with COVID-19, and the main change in imaging is ground-glass opacity. However, bilateral pneumonia is more common in adults, and the main change in imaging is also ground-glass opacity. (86, 87) One guideline (93) pointed out that there were multiple small patch shadows and interstitial changes in the early stages of the disease in adults, especially in the extrapulmonary zone in chest imaging. Furthermore, multiple ground-glass opacity or infiltrative shadows may develop in both lungs. In severe cases, pulmonary consolidation may occur, and pleural effusion is rare. An analysis of CT features in children with COVID-19 showed that in 15 cases, inflammatory infiltration was found in the chest CT imaging during initial diagnosis and reexamination. Most inflammatory infiltrations were manifested as small nodular ground-glass opacities, and multiple lobe segments were less involved. Multiple lobe segments were involved in only one case, and the imaging changes were not typical in the advanced stage as well (39). These findings suggest that pulmonary inflammation in children is mild and localized. The results of laboratory tests of children with COVID-19 were more often within the normal range than those of adults. The leukocyte count of children with COVID-19 was usually normal or below the normal range. The lymphocyte count was generally normal or above the normal range, and only 15% of cases were below the normal range. While for adults, the leukocyte count was usually normal or above the normal range, and the lymphocyte count were mostly below the normal range (35%-63%).(86, 87) It can be seen that there were significant differences in routine blood values between adults and children. However, some published COVID-19 guidelines and consensus for children consider a reduction in lymphocyte count as one of the factors for diagnosing suspected cases (90-92). Our study demonstrates that guidelines for children should not be formulated in full accordance with adult standards, otherwise true cases may be missed.

Most of the existing systematic reviews on characteristics of patients with COVID-19 are based on adult patients or patients regardless of age.(100-105) Two of these compared the differences between children and adult patients.(100, 101) Only one review reported the clinical characteristics of children with COVID-19 at present.(106) All reviews that considered children had similar outcomes: for example, children had milder symptoms than adults, some children had no symptoms, and lymphopenia in children did not occur as often as in adults. But our rapid review has included 49 studies of children patients, which is more than most of these reviews together. In contrast to another systematic review of children with COVID-19 (106), we included studies not only China but also from other countries such as Singapore, which were published in Chinese and English. Moreover, we conducted a meta-analysis whereas the previous study only did a systematic literature review.

### Strengths and limitations

This rapid review has several strengths. First, although this is not the first systemic review about the clinical characteristics of children with COVID-19, but is to our knowledge the first to combine the results with meta-analysis and GRADE evaluation of the quality of main evidence, which is of great importance for clinicians to diagnose and treat children rapidly. Second, our study points out the loopholes in some current guidance documents that suggest the diagnosis of suspected cases - also in children - based on the lymphocyte count. Third, as a rapid review, this study summarizes the latest published information on clinical cases, which provides relatively high-quality evidence for the formulation of clinical practice guidelines in the rapidly evolving public health emergency situation and helps policy-makers to make evidence-based decisions quickly (107).

Our study has also some limitations. First, due to the rapid fermentation of the public health emergency and new cases emerging continuously, the findings of this review may get outdates relatively soon. Second, cannot be sure if some cases were included in multiple studies. Third, at present, there is no unified definition for clinical classification of the severity of COVID-19, so we had to combine light, mild and moderate disease into one category (mild), while severe and critical cases were both considered as severe cases.

### Future implications

The researchers should aim to conduct more targeted studies on COVID-19 in specific subpopulations. Policy makers should develop accurate guidelines for both children and adults. Clinical practitioners should pay attention on the specific characteristics of different patient populations to improve the accuracy of diagnosis and treatment.

## Conclusion

Children with COVID-19 are more common to have only mild symptoms, and many children are even completely asymptomatic. Fever and cough are the main symptoms of COVID-19 in both children. Vomiting and diarrhea occurring less frequently in children. Ground-glass opacity is the most common CT imaging of children. Whereas adults tend to have elevated lymphocyte count at the beginning of the disease, in children the lymphocytes were usually within the normal range. As the characteristics of COVID-19 differ between adults and children in multiple ways, specific criteria for the diagnosis and treatment of COVID-19 in children are urgently needed.

## Data Availability

This rapid review is based on original researches,all data is from existing research and is true

## Author contributions

(I) Conception and design: E Liu, Y Chen; (II) Administrative support: E Liu, Y Chen; (III) Provision of study materials or patients: E Liu; (IV) Collection and assembly of data: Z Wang, Q Zhou, C Wang, Q Shi, S Lu, Y Xun; (V) Data analysis and interpretation: Z Wang, Q Zhou, C Wang, Y Ma, X Luo; (VI) Manuscript writing: All authors; (VII) Final approval of manuscript: All authors.

## Acknowledgments

We thank Janne Estill, Institute of Global Health of University of Geneva for providing guidance and comments for our review. We thank all the authors for their wonderful collaboration.

## Footnote

### Ethical Statement

The authors are accountable for all aspects of the work in ensuring that questions related to the accuracy or integrity of any part of the work are appropriately investigated and resolved.

## Supplementary Material

### Supplementary Material 1-Search strategy

#### PubMed

#1 “Epidemiological Studies”[Title/Abstract]

#2 “Epidemiologic* Study”[Title/Abstract]

#3 “Epidemiological characteristics”[Title/Abstract]

#4 “Clinical features”[Title/Abstract]

#5 “Clinical characteristics”[Title/Abstract]

#6 “Clinical Presentation”[Title/Abstract]

#7 #1-#6/ OR

#8 “COVID-19”[Supplementary Concept]

#9 “Severe Acute Respiratory Syndrome Coronavirus 2”[Supplementary Concept]

#10 “COVID-19”[Title/Abstract]

#11 “SARS-COV-2”[Title/Abstract]

#12 “Novel coronavirus” [Title/Abstract]

#13 “2019-novel coronavirus” [Title/Abstract]

#14 “coronavirus disease-19” [Title/Abstract]

#15 “coronavirus disease 2019” [Title/Abstract]

#16 “COVID19” [Title/Abstract]

#17 “Novel CoV” [Title/Abstract]

#18 “2019-nCoV” [Title/Abstract]

#19 “2019-CoV” [Title/Abstract]

#20 “Wuhan-Cov” [Title/Abstract]

#21 “Wuhan Coronavirus” [Title/Abstract]

#22 “Wuhan seafood market pneumonia virus” [Title/Abstract]

#23 #8-#22/ OR

#24 #7 AND #23

#### EMBASE

#1 ‘COVID-19’:ab,ti

#2 ‘SARS-COV-2’:ab,ti

#3 ‘novel coronavirus’:ab,ti

#4 ‘2019-novel coronavirus’:ab,ti

#5 ‘coronavirus disease-19’:ab,ti

#6 ‘coronavirus disease 2019’:ab,ti

#7 ‘COVID19’:ab,ti

#8 ‘novel cov’:ab,ti

#9 ‘2019-ncov’:ab,ti

#10 ‘2019-cov’:ab,ti

#11 ‘wuhan-cov’:ab,ti

#12 ‘wuhan coronavirus’:ab,ti

#13 ‘wuhan seafood market pneumonia virus’:ab,ti

#14 #1-13/OR

#15 ‘epidemiological studies’:ab,ti

#16 ‘epidemiologic* study’:ab,ti

#17 ‘epidemiological characteristics’:ab,ti

#18 ‘clinical feature’/exp

#19 ‘clinical features’:ab,ti

#20 ‘clinical characteristics’:ab,ti

#21 ‘clinical presentation’:ab,ti

#22 #15-21/OR

#23 #14 AND #22

#24 #23 AND [medline]/lim

#25 #23 NOT #24

#### Web of Science

#1 TOPIC: “Epidemiological Studies”

#2 TOPIC: “Epidemiologic* Study”

#3 TOPIC: “Epidemiological characteristics”

#4 TOPIC: “Clinical features”

#5 TOPIC: “Clinical characteristics”

#6 TOPIC: “Clinical Presentation”

#7 #1-6/OR

#8 TOPIC: “COVID-19”

#9 TOPIC: “SARS-COV-2”

#10 TOPIC: “Novel coronavirus”

#11 TOPIC: “2019-novel coronavirus”

#12 TOPIC: “coronavirus disease-19”

#13 TOPIC: “coronavirus disease 2019”

#14 TOPIC: “COVID19”

#15 TOPIC: “Novel CoV”

#16 TOPIC: “2019-nCoV”

#17 TOPIC: “2019-CoV”

#18 TOPIC: “Wuhan-Cov”

#19 TOPIC: “Wuhan Coronavirus”

#20 TOPIC: “Wuhan seafood market pneumonia virus”

#21 #8-20/ OR

#22 #7 AND #21

#### Cochrane

#1 “COVID-19”:ti,ab,kw

#2 “SARS-COV-2”:ti,ab,kw

#3 “Novel coronavirus”:ti,ab,kw

#4 “2019-novel coronavirus” :ti,ab,kw

#5 “Novel CoV” :ti,ab,kw

#6 “2019-nCoV” :ti,ab,kw

#7 “coronavirus disease-19” :ti,ab,kw

#8 “coronavirus disease 2019” :ti,ab,kw

#9 “COVID19” :ti,ab,kw

#10 “Wuhan-Cov” :ti,ab,kw

#11 “Wuhan Coronavirus” :ti,ab,kw

#12 “Wuhan seafood market pneumonia virus” :ti,ab,kw

#13 #1-12/OR

#14 “Epidemiological Studies”:ti,ab,kw

#15 “Epidemiologic* Study”:ti,ab,kw

#16 “Epidemiological characteristics”:ti,ab,kw

#17 “Clinical features”:ti,ab,kw

#18 “Clinical characteristics”:ti,ab,kw

#19 “Clinical Presentation”:ti,ab,kw

#20 #14-19/ OR

#21 #13 AND #20

#### CBM

#1 “新型冠状病毒”[常用字段:智能]

#2 “COVID-19”[常用字段:智能]

#3 “COVID 19”[常用字段:智能]

#4 “2019-nCoV”[常用字段:智能]

#5 “2019-CoV”[常用字段:智能]

#6 “SARS-CoV-2”[常用字段:智能]

#7 “武汉冠状病毒”[常用字段:智能]

#8 #1-7/OR

#9 “流行病学”[中文标题:智能]

#10 “临床表现”[中文标题:智能]

#11 “临床特征”[中文标题:智能]

#12 “临床特点”[中文标题:智能]

#13 #9-12/ OR

#14 #8 AND #13

#### WanFang

#1 “新型冠状病毒”[主题]

#2 “COVID-19”[主题]

#3 “COVID 19”[主题]

#4 “2019-nCoV”[主题]

#5 “2019-CoV”[主题]

#6 “SARS-CoV-2”[主题]

#7 “武汉冠状病毒”[主题]

#8 #1-#7/ OR

#9 “流行病学”[题名]

#10 “临床特点”[题名]

#11 “临床特征”[题名]

#12 “临床表现”[题名]

#13 #9-12/ OR

#14 #8 AND #13

#### CNKI

#1 “新型冠状病毒”[主题]

#2 “COVID-19”[主题]

#3 “COVID 19”[主题]

#4 “2019-nCoV”[主题]

#5 “2019-CoV”[主题]

#6 “SARS-CoV-2”[主题]

#7 “武汉冠状病毒”[主题]

#8 #1-7/ OR

#9 “流行病学”[篇名]

#10 “临床特点”[篇名]

#11 “临床特征”[篇名]

#12 “临床表现”[篇名]

#13 #9-12 OR

#14 #8 AND #13

### Supplementary Material 2-Risk of bias

**Table A.**
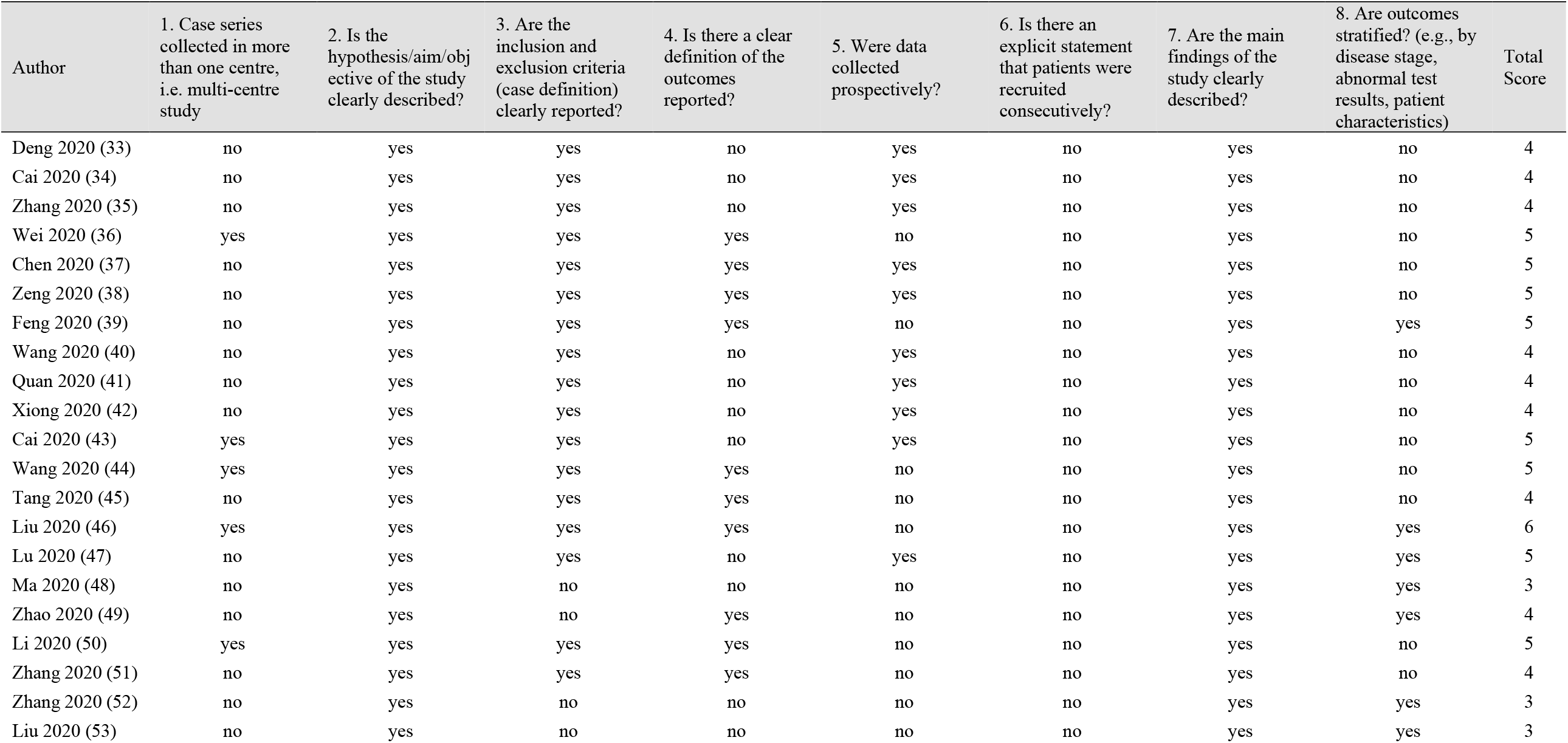
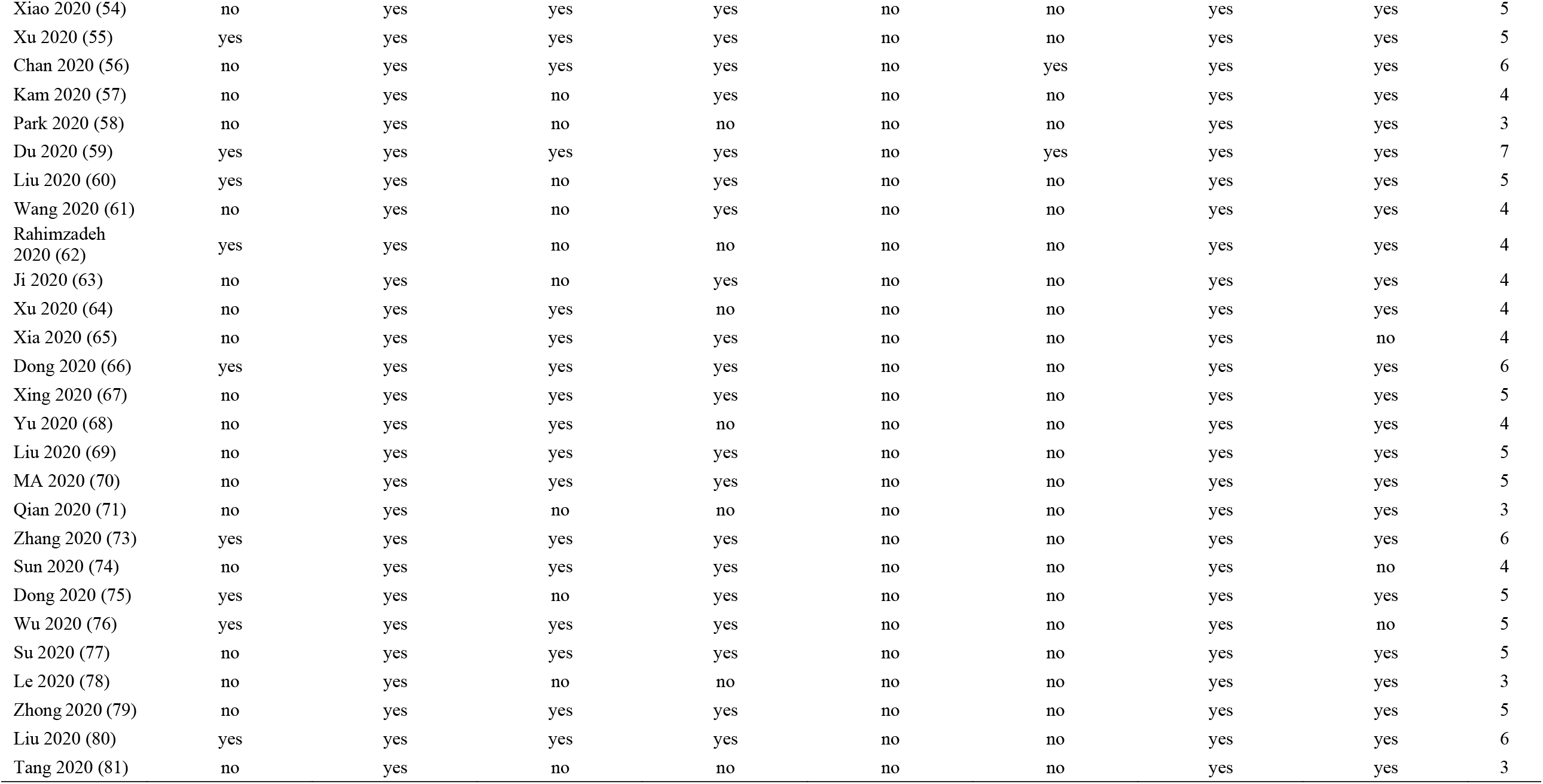
National Institute for Health and Care Excellence.

| Author | 1. Case series collected in more than one centre, i.e. multi-centre study | 2. Is the hypothesis/aim/objective of the study clearly described? | 3. Are the inclusion and exclusion criteria (case definition) clearly reported? | 4. Is there a clear definition of the outcomes reported? | 5. Were data collected prospectively? | 6. Is there an explicit statement that patients were recruited consecutively? | 7. Are the main findings of the study clearly described? | 8. Are outcomes stratified? (e.g., by disease stage, abnormal test results, patient characteristics) | Total Score |
| --- | --- | --- | --- | --- | --- | --- | --- | --- | --- |
| Deng 2020 (33) | no | yes | yes | no | yes | no | yes | no | 4 |
| Cai 2020 (34) | no | yes | yes | no | yes | no | yes | no | 4 |
| Zhang 2020 (35) | no | yes | yes | no | yes | no | yes | no | 4 |
| Wei 2020 (36) | yes | yes | yes | yes | no | no | yes | no | 5 |
| Chen 2020 (37) | no | yes | yes | yes | yes | no | yes | no | 5 |
| Zeng 2020 (38) | no | yes | yes | yes | yes | no | yes | no | 5 |
| Feng 2020 (39) | no | yes | yes | yes | no | no | yes | yes | 5 |
| Wang 2020 (40) | no | yes | yes | no | yes | no | yes | no | 4 |
| Quan 2020 (41) | no | yes | yes | no | yes | no | yes | no | 4 |
| Xiong 2020 (42) | no | yes | yes | no | yes | no | yes | no | 4 |
| Cai 2020 (43) | yes | yes | yes | no | yes | no | yes | no | 5 |
| Wang 2020 (44) | yes | yes | yes | yes | no | no | yes | no | 5 |
| Tang 2020 (45) | no | yes | yes | yes | no | no | yes | no | 4 |
| Liu 2020 (46) | yes | yes | yes | yes | no | no | yes | yes | 6 |
| Lu 2020 (47) | no | yes | yes | no | yes | no | yes | yes | 5 |
| Ma 2020 (48) | no | yes | no | no | no | no | yes | yes | 3 |
| Zhao 2020 (49) | no | yes | no | yes | no | no | yes | yes | 4 |
| Li 2020 (50) | yes | yes | yes | yes | no | no | yes | no | 5 |
| Zhang 2020 (51) | no | yes | yes | yes | no | no | yes | no | 4 |
| Zhang 2020 (52) | no | yes | no | no | no | no | yes | yes | 3 |
| Liu 2020 (53) | no | yes | no | no | no | no | yes | yes | 3 |
| Xiao 2020 (54) | no | yes | yes | yes | no | no | yes | yes | 5 |
| Xu 2020 (55) | yes | yes | yes | yes | no | no | yes | yes | 5 |
| Chan 2020 (56) | no | yes | yes | yes | no | yes | yes | yes | 6 |
| Kam 2020 (57) | no | yes | no | yes | no | no | yes | yes | 4 |
| Park 2020 (58) | no | yes | no | no | no | no | yes | yes | 3 |
| Du 2020 (59) | yes | yes | yes | yes | no | yes | yes | yes | 7 |
| Liu 2020 (60) | yes | yes | no | yes | no | no | yes | yes | 5 |
| Wang 2020 (61) | no | yes | no | yes | no | no | yes | yes | 4 |
| Rahimzadeh<br>2020 (62) | yes | yes | no | no | no | no | yes | yes | 4 |
| Ji 2020 (63) | no | yes | no | yes | no | no | yes | yes | 4 |
| Xu 2020 (64) | no | yes | yes | no | no | no | yes | yes | 4 |
| Xia 2020 (65) | no | yes | yes | yes | no | no | yes | no | 4 |
| Dong 2020 (66) | yes | yes | yes | yes | no | no | yes | yes | 6 |
| Xing 2020 (67) | no | yes | yes | yes | no | no | yes | yes | 5 |
| Yu 2020 (68) | no | yes | yes | no | no | no | yes | yes | 4 |
| Liu 2020 (69) | no | yes | yes | yes | no | no | yes | yes | 5 |
| MA 2020 (70) | no | yes | yes | yes | no | no | yes | yes | 5 |
| Qian 2020 (71) | no | yes | no | no | no | no | yes | yes | 3 |
| Zhang 2020 (73) | yes | yes | yes | yes | no | no | yes | yes | 6 |
| Sun 2020 (74) | no | yes | yes | yes | no | no | yes | no | 4 |
| Dong 2020 (75) | yes | yes | no | yes | no | no | yes | yes | 5 |
| Wu 2020 (76) | yes | yes | yes | yes | no | no | yes | no | 5 |
| Su 2020 (77) | no | yes | yes | yes | no | no | yes | yes | 5 |
| Le 2020 (78) | no | yes | no | no | no | no | yes | yes | 3 |
| Zhong 2020 (79) | no | yes | yes | yes | no | no | yes | yes | 5 |
| Liu 2020 (80) | yes | yes | yes | yes | no | no | yes | yes | 6 |
| Tang 2020 (81) | no | yes | no | no | no | no | yes | yes | 3 |

**Table B.** Newcastle-Ottawa Scale.

| Author | SELECTION |  |  | COMPARABILITY |  | OUTCOME |  |  |
| --- | --- | --- | --- | --- | --- | --- | --- | --- |
|  | 1) Representativeness of the Exposed Cohort | 2) Selection of the Non-Exposed Cohort | 3) Ascertainment of Exposure | 4) Demonstration That Outcome of Interest Was Not Present at Start of Study | 1) Comparability of Cohorts on the Basis of the Design or Analysis | 1) Assessment of Outcome | 2) Was Follow-Up Long Enough for Outcomes to Occur | 3) Adequacy of Follow Up of Cohorts |
| Qiu<br>2020<br>(72) | * |  | * | * |  | * |  |  |

### Supplementary Material 3-GRADE evidence profile

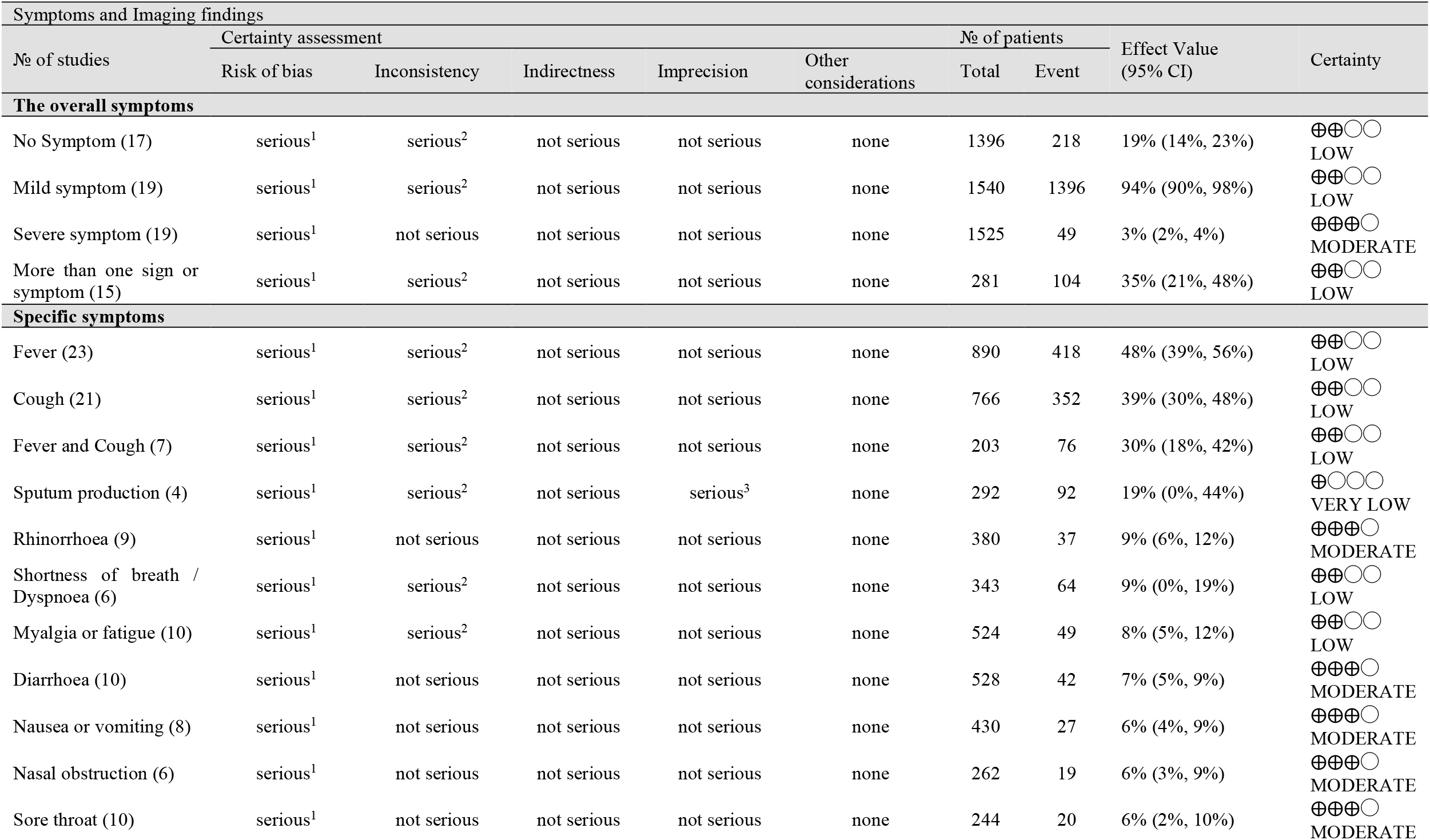
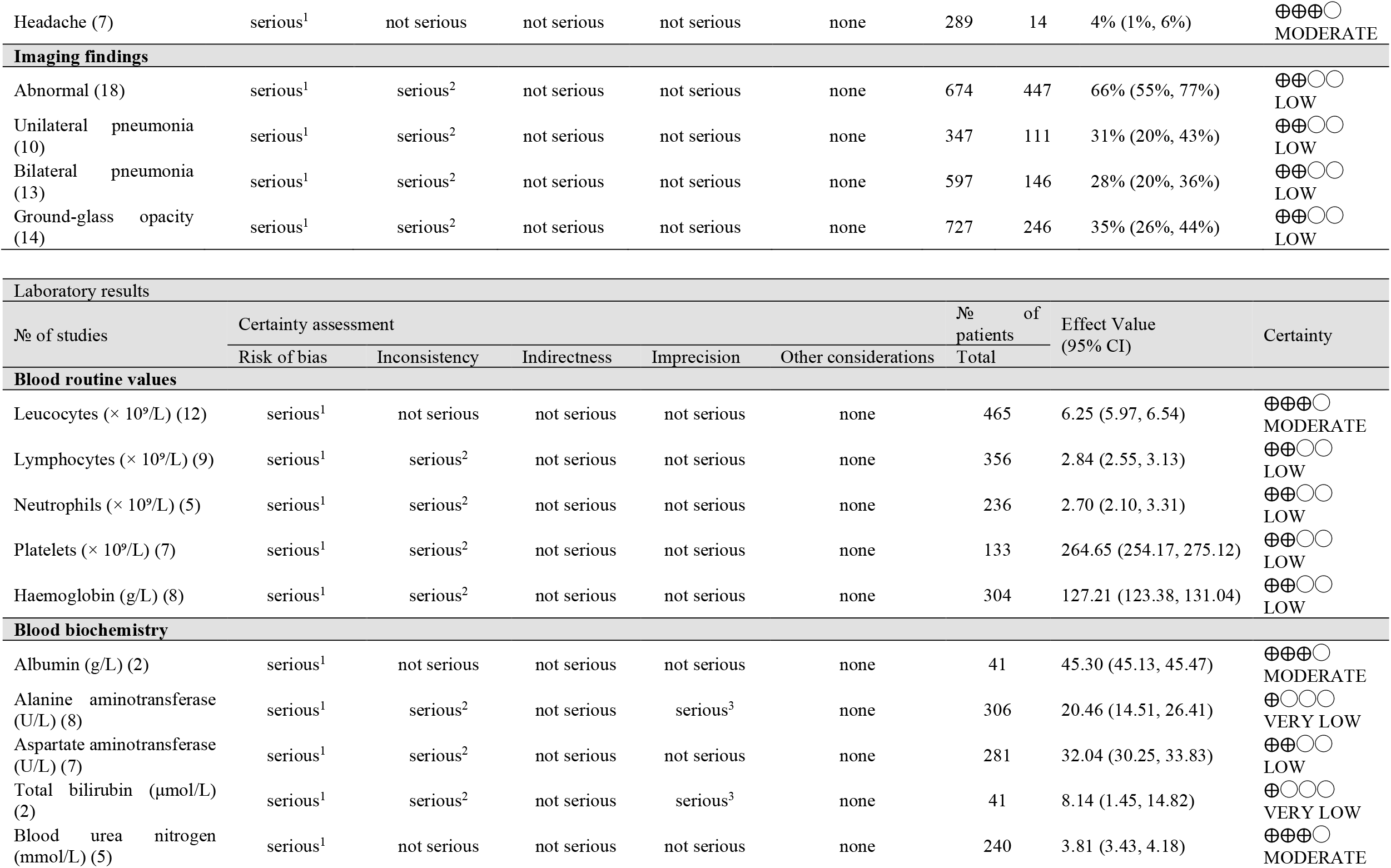
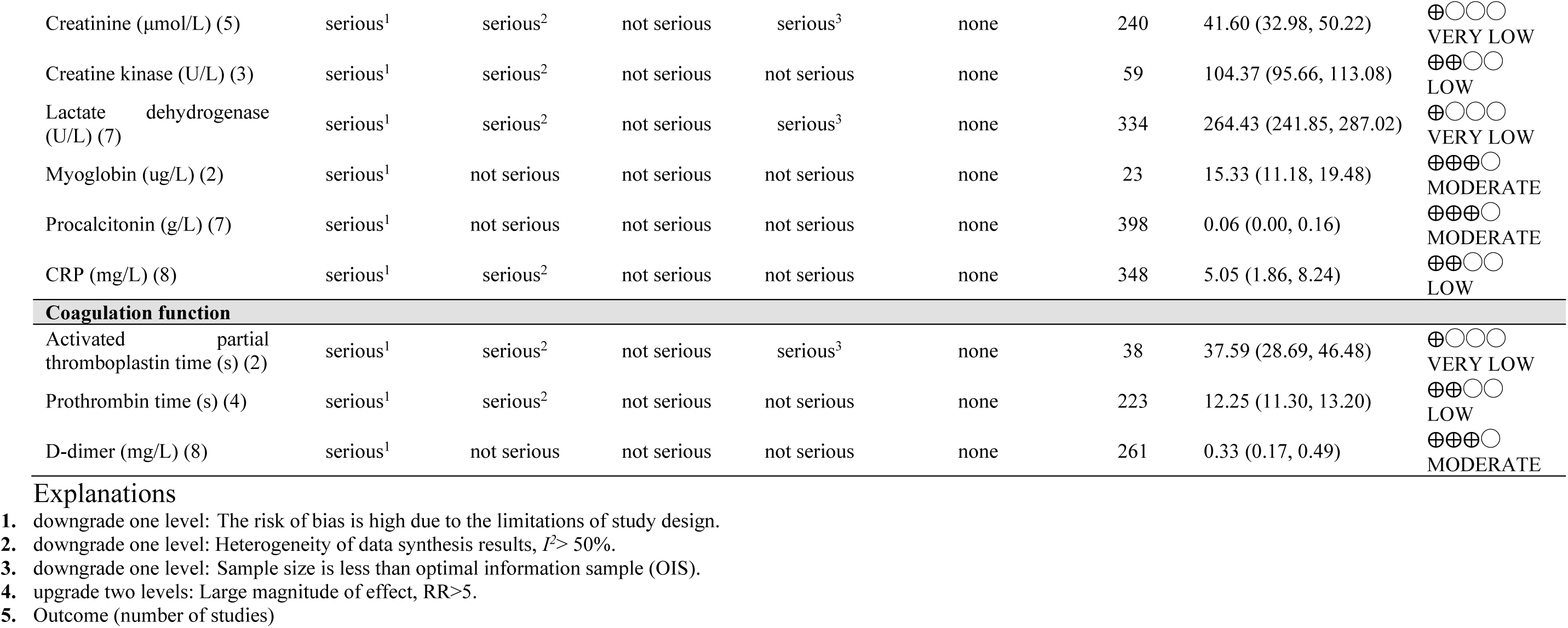

### Supplementary Material 4-Case report summary

**Table A.**
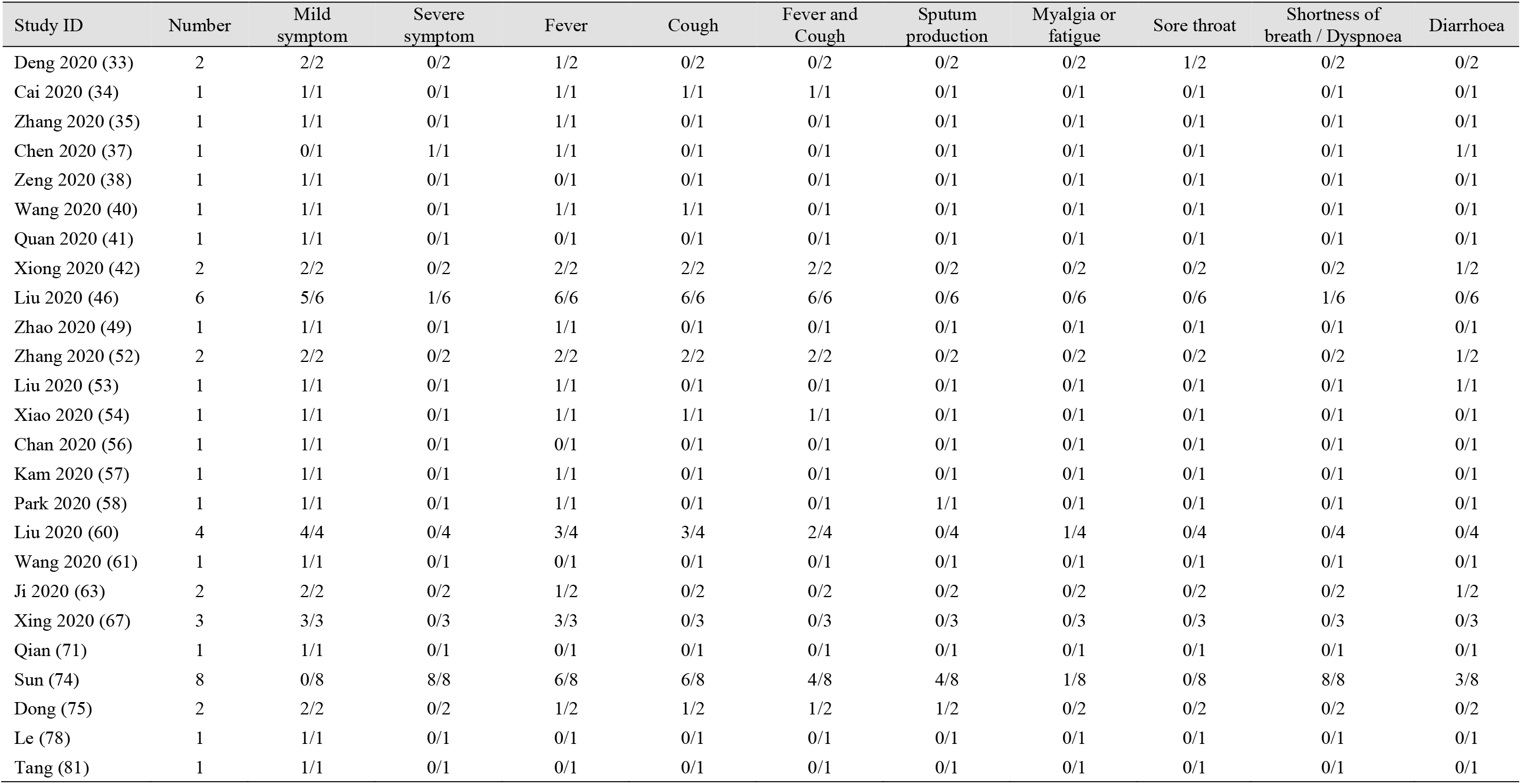
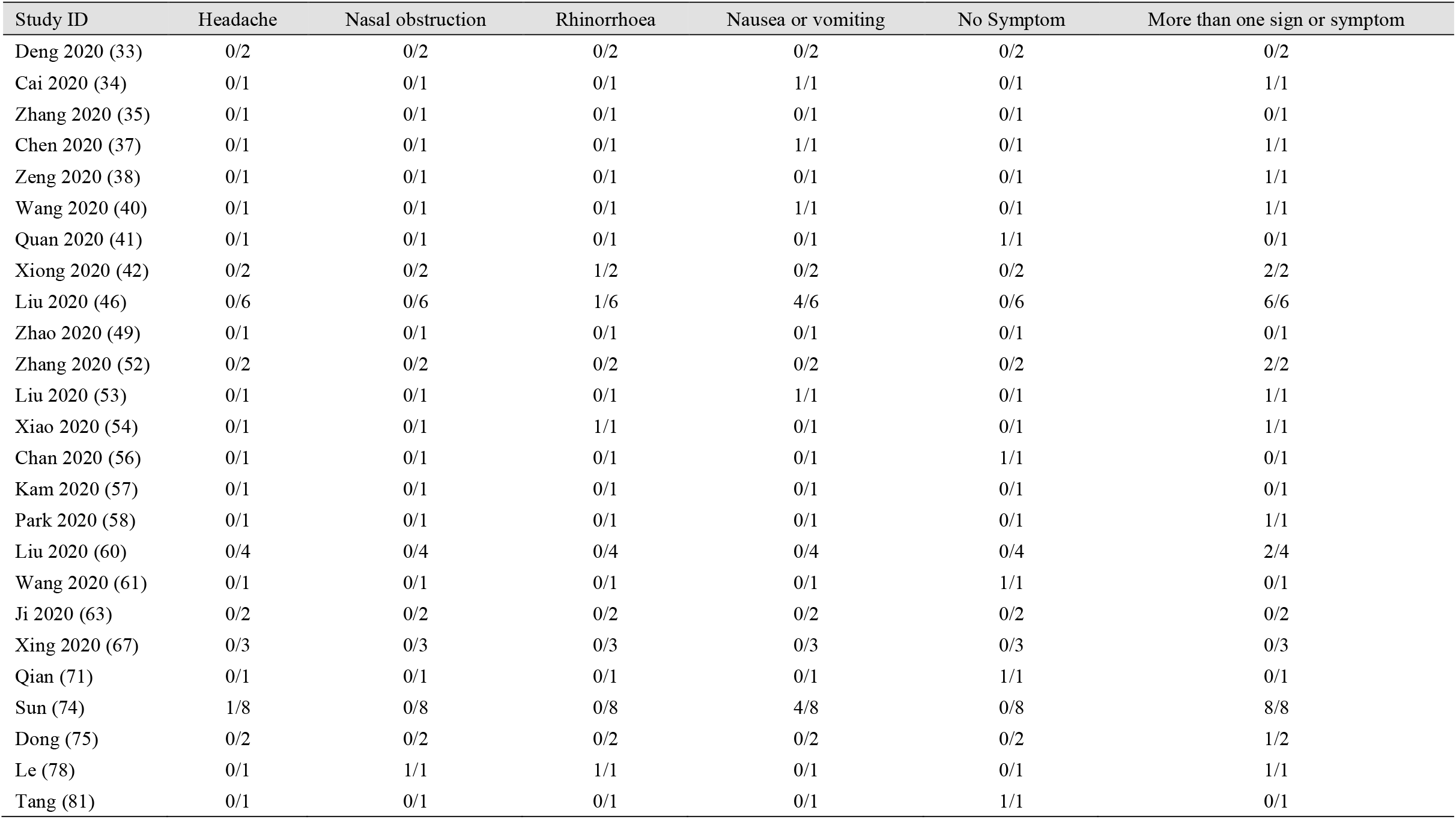
Symptom.

**Table B.** Imaging findings and Family contact.

| Study ID | Number | Abnormal | Unilateral pneumonia | Bilateral pneumonia | Ground-glass opacity | Family contact |
| --- | --- | --- | --- | --- | --- | --- |
| Deng 2020 (33) | 2 | 0/2 | 0/2 | 0/2 | 0/2 | 2/2 |
| Cai 2020 (34) | 1 | 1/1 | NR/1 | NR/1 | NR/1 | 1/1 |
| Zhang 2020 (35) | 1 | 1/1 | NR/1 | NR/1 | NR/1 | NR/1 |
| Chen 2020 (37) | 1 | 1/1 | 1/1 | 0/1 | 1/1 | 0/1 |
| Zeng 2020 (38) | 1 | 1/1 | 0/1 | 1/1 | 0/1 | 1/1 |
| Wang 2020 (40) | 1 | 1/1 | 1/1 | 0/1 | 1/1 | 1/1 |
| Quan 2020 (41) | 1 | 1/1 | 1/1 | 0/1 | 0/1 | 1/1 |
| Xiong 2020 (42) | 2 | 2/2 | 2/2 | 0/2 | 1/2 | 2/2 |
| Liu 2020 (46) | 6 | 4/5 | 0/5 | 4/5 | 1/5 | 0/6 |
| Zhao 2020 (49) | 1 | 1/1 | 1/1 | 0/1 | 0/1 | 1/1 |
| Zhang 2020 (52) | 2 | 1/2 | 0/2 | 1/2 | NR/2 | 2/2 |
| Liu 2020 (53) | 1 | 1/1 | 0/1 | 1/1 | 1/1 | 1/1 |
| Xiao 2020 (54) | 1 | 1/1 | 0/1 | 1/1 | 0/1 | 1/1 |
| Chan 2020 (56) | 1 | 1/1 | NR/1 | NR/1 | 1/1 | 1/1 |
| Kam 2020 (57) | 1 | NR/1 | NR/1 | NR/1 | NR/1 | 1/1 |
| Park 2020 (58) | 1 | 1/1 | 1/1 | 0/1 | 1/1 | 1/1 |
| Liu 2020 (60) | 4 | 3/4 | NR/4 | NR/4 | 1/4 | NR/4 |
| Wang 2020 (61) | 1 | 1/1 | NR/1 | NR/1 | NR/1 | 1/1 |
| Ji 2020 (63) | 2 | 0/2 | 0/2 | 0/2 | 0/2 | 2/2 |
| Xing 2020 (67) | 3 | 2/3 | 2/3 | 0/3 | 2/3 | 3/3 |
| Qian (71) | 1 | NR/1 | NR/1 | NR/1 | NR/1 | 1/1 |
| Sun (74) | 8 | 8/8 | 2/8 | 6/8 | 6/8 | 5/8 |
| Dong (75) | 2 | 1/2 | 0/2 | 1/2 | NR/2 | NR/2 |
| Le (78) | 1 | 0/1 | 0/1 | 0/1 | 0/1 | 1/1 |
| Tang (81) | 1 | 0/1 | 0/1 | 0/1 | 0/1 | 0/1 |

NR: not report

**Table C.** Laboratory results.

| Study ID | Number | Leucocytes ($\times 10^9/L$) | Lymphocytes ($\times 10^9/L$) | Neutrophils ($\times 10^9/L$) | Platelets ($\times 10^9/L$) | Haemoglobin (g/L) |
| --- | --- | --- | --- | --- | --- | --- |
| Deng 2020 (33) | 2 | 4.83/5.08 | 3.37/2.89 | NR | 225/287 | NR |
| Cai 2020 (34) | 1 | 16.0 | NR | NR | 138 | NR |
| Zhang 2020 (35) | 1 | 9.68 | NR | NR | 494 | 113 |
| Chen 2020 (37) | 1 | 7.52 | NR | NR | 183 | 108 |
| Zeng 2020 (38) | 1 | 7.66 | NR | NR | 399 | 132 |
| Wang 2020 (40) | 1 | 16.09 | 1.38 | 13.97 | 278 | NR |
| Quan 2020 (41) | 1 | 9.1 | 4.5 | NR | 234 | 138 |
| Xiong 2020 (42) | 2 | 6.0/6.2 | 2.56/3.73 | 2.18/1.72 | 301/252 | 112/117 |
| Liu 2020 (46) | 6 | 2.96/6.49/5.48/3.04/3.95/1.8 | 1/1.19/1.25/1.7/0.87/0.36 | 1.3/3.15/2.54/0.66/2.85/0.27 | 203/272/256/191/153/165 | 104/120/113/118/115/120 |
| Zhao 2020 (49) | 1 | 3.9 | NR | NR | NR | NR |
| Zhang 2020 (52) | 2 | 5.94/8.98 | NR | NR | 255/324 | 111/112 |
| Liu 2020 (53) | 1 | 3.78 | 1.86 | 1.68 | 149 | 138 |
| Xiao 2020 (54) | 1 | Normal | Normal | Normal | Normal | Normal |
| Chan 2020 (56) | 1 | 6.5 | 2.8 | 3.2 | 197 | 146 |
| Kam 2020 (57) | 1 | Normal | Normal | Normal | Normal | Normal |
| Park 2020 (58) | 1 | 4.08 | NR | NR | 251 | 135 |
| Liu 2020 (60) | 4 | Decreased/Normal/Normal/Normal | Increased/Increased/Normal/Normal | NR | NR | NR |
| Wang 2020 (61) | 1 | NR | 2.43 | NR | NR | NR |
| Ji 2020 (63) | 2 | 11.82/6.6 | NR | NR | NR | NR |
| Xing 2020 (67) | 3 | 7.3/9.6/6.0 | 5.4/5.2/4.9 | 1.2/3.6/1.7 | 333.0/411.0/186.0 | 332.0/359.0/332.0 |
| Qian (71) | 1 | NR | NR | NR | NR | NR |
| Sun (74) | 8 | 1.65/14.95/9.19/8.32/8.8/10.6/3.85/7.6 | 0.69/1.96/2.7/6.41/3.6/4.04/1.7/2.8 | 0.78/11.63/5.7/1.27/3.5/5.9/1.9/3.8 | 140/68/145/666/247/515/154/250 | 83/90/103/111/123/150/159/136 |
| Dong (75) | 2 | NR/normal | NR/normal | NR/normal | NR/normal | NR/normal |
| Le (78) | 1 | 10.23 | 8.3 | 1.19 | 230 | 112 |
| Tang (81) | 1 | NR | NR | NR | NR | NR |

**continue**
| Study ID | Albumin (g/L) | Alanine aminotransferase (U/L) | Aspartate aminotransferase (U/L) | Total bilirubin (μmol/L) | Blood urea nitrogen (mmol/L) | Creatinine (μmol/L) |
| --- | --- | --- | --- | --- | --- | --- |
| Deng 2020 (33) | NR | 10.63/12.25 | 23.32/20.87 | NR | NR | 27.33/35.29 |
| Cai 2020 (34) | NR | 17 | 33 | NR | NR | 29 |
| Zhang 2020 (35) | NR | NR | NR | NR | NR | NR |
| Chen 2020 (37) | Normal | Normal | Normal | Normal | 15.9 | 224 |
| Zeng 2020 (38) | Normal | Normal | Normal | Normal | Normal | Normal |
| Wang 2020 (40) | Normal | 20 | 31 | Normal | Normal | Normal |
| Quan 2020 (41) | Normal | Normal | Normal | Normal | Normal | Normal |
| Xiong 2020 (42) | 43.2/Normal | 58.5/Normal | 83.5/Normal | NR/Normal | Normal | Normal |
| Liu 2020 (46) | 40/45.2/43.6/45.4/44.3/42.3 | 6/14/11/23/43/15 | 45/30/42/64/36/37 | NR/4/4.2/2.7/3.6/5.4 | NR | 33/34/22/29/23/30 |
| Zhao 2020 (49) | Normal | Normal | Normal | Normal | 496 | Normal |
| Zhang 2020 (52) | NR/NR | 12.48/13.11 | 45.48/63.94 | NR | 5.16/5.30 | 21.6/17.90 |
| Liu 2020 (53) | NR | NR | NR | NR | NR | NR |
| Xiao 2020 (54) | Normal | Normal | Normal | Normal | Normal | Normal |
| Chan 2020 (56) | 49.1 | 23.9 | 28.2 | 3.6 | 5.6 | 51 |
| Kam 2020 (57) | NR | NR | NR | NR | NR | NR |
| Park 2020 (58) | NR | NR | NR | NR | NR | NR |
| Liu 2020 (60) | NR | NR | NR | NR | NR | NR |
| Wang 2020 (61) | NR | NR | 143 | 33 | NR | NR |
| Ji 2020 (63) | NR | NR | NR | NR | NR | NR |
| Xing 2020 (67) | NR | NR | NR | NR | NR | 22.8/28.3/53.4 |
| Qian (71) | NR | NR | NR | NR | NR | NR |
| Sun (74) | NR | 58/66/36/100/55/9/16/8 | 37/27/33/41/16/14/14/16 | 11.8/20.4/16.5/12.4/5.3/7.8/8.1/8.1 | NR | 27.1/43.4/21.3/15/24.8/64.5/58/72.1 |
| Dong (75) | NR/normal | NR/normal | NR/normal | NR/normal | NR/normal | NR/normal |
| Le (78) | NR | 34.8 | 59.9 | NR | NR | 36.5 |
| Tang (81) | NR | NR | NR | NR | NR | NR |

continue
| Study ID | Creatine kinase (U/L) | Lactate dehydrogenase (U/L) | Myoglobin (ug/L) | Procalcitonin (ng/mL) | CRP (mg/L) | Activated partial thromboplastin time (s) | Prothrombin time (s) | D-dimer (mg/L) |
| --- | --- | --- | --- | --- | --- | --- | --- | --- |
| Deng 2020 (33) | 75.33/83.78 | NR | NR | 0.19/<0.05 | 12/<10 | Normal/NR | Normal/NR | 0.41/NR |
| Cai 2020 (34) | NR | NR | NR | 0.07 | 15 | Normal | Normal | 0.58 |
| Zhang 2020 (35) | NR | NR | NR | 0.073 | 5.66 | NR | NR | NR |
| Chen 2020 (37) | NR | NR | NR | NR | NR | NR | 14.3 | Normal |
| Zeng 2020 (38) | NR | NR | NR | 0.08 | < 0.75 | NR | NR | NR |
| Wang 2020 (40) | NR | NR | NR | NR | NR | Normal | Normal | Normal |
| Quan 2020 (41) | Normal | Normal | Normal | NR | <0.499 | NR | NR | NR |
| Xiong 2020 (42) | NR/Normal | NR | NR | NR/0.05 | 0.36/7.81 | Normal | Normal | Normal |
| Liu 2020 (46) | 29/50/77/148/71/82 | 384/197/476/375/297/280 | NR | NR | 38.4/21/23.32/11.8/58.79/6.84 | NA/32.2/43.7/41/34/41.5 | NR/13.2/12.8/11.9/12.3/12.5 | 0.59/0.22/0.78/0.22/0.74/0.38 |
| Zhao 2020 (49) | Normal | Normal | Normal | 0.06 | Normal | NR | NR | NR |
| Zhang 2020 (52) | 30.48/102.53 | 318.37/540.33 | NR | 0.85/0.23 | 0.05/0.04 | Normal | Normal | Normal |
| Liu 2020 (53) | NR | NR | NR | NR | NR | NR | NR | NR |
| Xiao 2020 (54) | Normal | Normal | Normal | Normal | Normal | Normal | Normal | Normal |
| Chan 2020 (56) | 78 | 194 | NR | NR | 0.2 | 34.0 | 13.1 | NR |
| Kam 2020 (57) | NR | NR | NR | NR | NR | NR | NR | NR |
| Park 2020 (58) | NR | NR | NR | NR | < 0.04 | NR | NR | NR |
| Liu 2020 (60) | NR | NR | NR | NR | Normal/Normal/Normal/Increased | NR | NR | NR |
| Wang 2020 (61) | 479 | NR | NR | NR | NR | NR | NR | NR |
| Ji 2020 (63) | NR | NR | NR | NR | 34.64/3.49 | NR | NR | NR |
| Xing 2020 (67) | 73.2/88.6/91.0 | 264.3/NR/194.0 | NR | 0.23/0.21/0.73 | <0.8/<5.0/10.5 | NR | NR | 860.0/230.0/190.0 |
| Qian (71) | NR | NR | NR | NR | NR | NR | NR | NR |
| Sun (74) | 15/20,702/33/148/262/106/72/77 | 394/888/282/891/471/370/209/187 | NR | 0.18/17.16/0.05/0.08/0.11/0.04/0.09/0.05 | 6.48/57.9/103/0.75/27.02/1/9.9/0.5 | NR | NR | 0.47/40.34/3.07/NR/NR/NR/0.23/0.44 |
| Dong (75) | NR/normal | NR/normal | NR/normal | NR/normal | NR/normal | NR/normal | NR/normal | NR/normal |
| Le (78) | NR | 327 | NR | 0.08 | 0.32 | NR | NR | NR |
| Tang (81) | NR | NR | NR | NR | NR | NR | NR | NR |

NR: not report; NA: not applicable

